# Identifying and Characterising Common Genetic Differences in Schizophrenia and Bipolar Disorder

**DOI:** 10.64898/2026.07.17.26358311

**Authors:** Isabella R. Willcocks, Alex Richards, Sophie E. Legge, Peter Holmans, Arianna Di Florio, Alastair G. Cardno, Michael C. O’Donovan, Michael J. Owen, Antonio F. Pardiñas, James T. R. Walters

## Abstract

Schizophrenia and bipolar disorder are diagnostically distinct categories that overlap substantially in clinical features and genetic aetiology. Understanding genetic variants that contribute liability specifically to each disorder can offer insights into biological processes that differentiate them. Here we used Case-Case GWAS (CC-GWAS) to identify common genetic variants differentially associated with schizophrenia and bipolar disorder, analysing 67,390 schizophrenia cases and 41,917 bipolar disorder cases. We identified 19 genome-wide significant loci, of which 16 (84%) demonstrated divergent genetic effects with risk alleles showing opposite directions of association between disorders. The CC-GWAS summary statistics had detectable disorder-differentiating heritability (10.27%, SE=0.01) and showed genetic correlations indicating that SCZ-differentiating alleles were associated with lower educational attainment, lower cognitive performance, and increased risk of ADHD, anorexia, autism, BD1 (though not BD2), cannabis use disorder, and OCD. Four loci showed divergent effects despite not reaching genome-wide significance in either individual disorder GWAS, demonstrating enhanced power to detect opposite-direction effects. Functional annotation identified 102 mapped genes significantly enriched for expression across all 13 tested brain regions, with no significant enrichment in peripheral tissues, and gene set enrichment analysis implicated neuronal projection and synaptic compartments as the strongest biological themes differentiating the two disorders. Polygenic risk scores derived from these disorder-differentiating variants were associated with earlier age at onset and more severe negative symptoms in schizophrenia, consistent with these variants marking neurodevelopmental dimensions of illness. Our findings provide targets for understanding pathogenic differences between schizophrenia and bipolar disorder and demonstrate that genuine divergent genetic effects exist beyond the substantial shared liability.

## Introduction

Schizophrenia (SCZ) and bipolar disorder (BD) can be severe, often lifelong, psychiatric illnesses that share symptoms and associated features such as psychosis, mania, depression and cognitive impairment^1^. Their clinical presentations and courses are variable, but both disorders can lead to persistent disability and constitute significant public health concerns. For decades, genetic research has attempted to identify and delimit the biological basis of these disorders with the aim of improving their diagnosis and treatment. Classic genetic epidemiology (for example twin studies) has consistently shown SCZ and BD to have the highest heritability metrics of common adult-onset psychiatric conditions, attributing 60-80% of the variance in liability to genetic factors^2–4^. A large proportion of the genetic susceptibility is driven by variants shared between the two conditions, with observations from genetic epidemiology^5,6^ and from recent large-scale genome-wide association studies (GWAS) showing that the two conditions have the highest genetic correlation (0.68) of any pair of common psychiatric diagnoses^7^. Genetic variants that confer susceptibility to both SCZ and BD could highlight common molecular pathways implicated in the broad spectrum of psychotic diagnoses, an area of research that has been recently reviewed^8^. Alternatively, finding specific risk alleles that differentiate these conditions could lead to the identification of biological processes specific to their clinical presentations, which could inform biomarker identification to assist patient stratification, inform clinical subtyping, and ultimately identify novel therapeutics^9^.

Genomic studies have attempted to investigate how transdiagnostic symptoms and associated clinical features between SCZ and BD relate to polygenic liability for these and other psychiatric disorders^10^. This is particularly relevant given that the diagnostic boundaries between these disorders are often unclear. For instance, schizoaffective disorder (bipolar subtype) occupies an intermediate position between SCZ and BD and shares features of both. Consistent with this, liability to SCZ, as indexed by polygenic risk scores (PRS), has been shown to differ between those with schizoaffective disorder (bipolar subtype) and those with other diagnoses of BD^11,12^. Further work using exploratory and confirmatory factor analyses identified statistically distinct symptom dimensions within people with BD (psychosis, mania and depression) and showed that major depression (MDD) PRS was specifically associated with the depression dimension and the SCZ PRS with the psychosis dimension^13^. Finally, applying a structural equation modelling framework (Genomic SEM) to GWAS summary statistics for BD, SCZ and MDD identified latent ‘differentiating’ components unique to each disorder, as well as a common ‘shared’ component^14^. PRS derived from these differentiating components were then shown to be differentially associated with dimensional symptom scores in people with BD, with the components for BD, SCZ and MDD respectively associating with dimensions of mania, mood-incongruent psychosis, and depression^14^.

Another approach to identifying alleles that differentiate SCZ from BD is to conduct so called “case-case” analyses. The largest of these (23,585 SCZ cases and 15,270 BD cases) identified 2 genome - wide significant (GWS) loci with divergent effects on SCZ and BD; rs56355601 within an intron of DARS2, and rs200005157 within an intron of ARFGEF2^15^. That study, as well as efforts on smaller samples ^16,17^ relied on access to individual-level genomic data, which are not usually publicly available and require strict harmonisation protocols to avoid confounding, for example from ancestral/population stratification and genotyping array/batch effects^18^. Methods that allow the use of GWAS summary statistics, which are generally more widely available, allow genetic differences between disorders to be investigated when it is not feasible to perform individual-level case-case analyses. Indeed, a recently published method for jointly analysing summary data from two case-control GWAS, CC-GWAS^19^, identified 12 significant loci when applied to SCZ (40,675 cases, 64,643 controls)^20^ and BD (20,352 cases, 31,358 controls)^21^, using the largest available GWAS at the time of publication.

In the current study, we apply CC-GWAS to recent studies of SCZ and BD, increasing the case sample size by 65% in SCZ and 105% in BD, to identify loci specific to either disorder. After refining the detected loci using GenomicSEM to identify regions with evidence of pleiotropy, we performed a series of downstream analyses on the CC-GWAS summary statistics, including polygenic risk scores, heritability estimates and genetic correlations. These downstream analyses allow us to demonstrate the utility of CC-GWAS beyond locus discovery, offering a route to understanding the biological processes that differentiate SCZ from BD.

## Methods

Statistical analyses, data curation and data visualisations were, unless otherwise specified, completed in R (v4.5.2) via RStudio (2026.01.0 Build 392).

### Source GWAS

For the SCZ GWAS, we used the most recent study conducted by the Schizophrenia Working Group of the Psychiatric Genomics Consortium (PGC)^22^. The results discussed here are based upon the combined analysis of the European and East Asian PGC cohorts (the “core PGC dataset”). A sensitivity analysis showed that restricting our CC-GWAS to summary statistics derived solely from European samples did not significantly alter the results, with a genetic correlation of Rg=1 (SE=0.0028, P=2.52E-27266) between the cross-ancestry and European-only CC-GWAS results. The total sample size was 67,390 cases (a combination of schizophrenia and schizoaffective disorder), 94,015 controls and the dataset included 7,585,076 SNPs, imputed using the Haplotype Reference Consortium (HRC) release 1.1^23^. Detailed information can be found elsewhere^22^.

The 2021 GWAS published by the Bipolar Disorder Working Group of the PGC was used in the current analysis^24^. While a larger BD GWAS was recently published^25^, we opted to use the 2021 GWAS due to its narrower phenotypic definition. The 2025 study’s substantial increase in sample size primarily derives from self-reported diagnoses, and the authors report significant differences in genetic architecture between clinically ascertained and self-reported cases. Given the well-documented challenges with diagnostic accuracy in self-reported bipolar disorder, with studies showing that fewer than half of individuals who self-report a bipolar disorder diagnosis meet criteria on structured clinical interviews^26^, we prioritised phenotypic specificity over sample size for our analyses. The BD GWAS contained a total of 57 cohorts, primarily of European ancestry. The summary statistics were generated based on 41,917 cases, a combination of BD type 1 and type 2 (justification for using combined summary statistics provided in **Supplementary Table 1**), 371,549 controls and 7,825,140 SNPs, also imputed against the HRC 1.1 panel^23^. Detailed information has been provided previously^24^.

### CC-GWAS

CC-GWAS requires six parameters: case and control counts, degree of control overlap between the samples, lifetime disorder prevalence (K), SNP-based heritability on the liability scale, genetic correlation between the disorders (Rg) and the estimated number of independently associated causal SNPs (m). Control overlap can boost the power of the CC-GWAS method by allowing for a more direct comparison of the case cohorts^19^.

Using CC-GWAS terminology, SCZ was designated as “disorder A” and BD as “disorder B”; positive betas therefore indicate the specified allele confers greater risk for SCZ relative to BD. The lifetime disorder prevalences and SNP-based heritabilities (liability scale) were taken directly from the source GWAS publications (K =1% and h^2^ =0.18 for SCZ^22^, and K =2% and h^2^ =0.19 for BD^24^). We estimated Rg=0.68 using LD score regression^27^. As recommended by the method developers^19^, a parameter representing the number of independent causal SNPs for the two disorders was taken as the average number of these reported in independent studies^28^ (8300 and 6400 SNPs for SCZ and BD respectively, yielding m=7350). Control overlap between the two GWAS was 45,497 individuals, as provided by analysts from the PGC Schizophrenia Working Group.

For inclusion in CC-GWAS, variants required a minor allele frequency (MAF) ≥ 0.01 and a meta-analysed sample size for that SNP exceeding two thirds of the maximum possible effective sample size (Neff)^29^. This led to retaining 7,394,639 SNPs for SCZ and 7,590,773 SNPs for BD, with 7,341,513 variants available in both datasets. As expected, given that the same imputation platform and quality control pipeline were used for both GWAS^30^, there were no discordant base pair or chromosome positions, and all reference alleles were aligned correctly between both datasets. Following removal of strand-ambiguous SNPs (A/T and C/G), 6,229,286 SNPs were available for CC-GWAS.

### LD-based Clumping of Loci

LD-based clumping of loci was conducted via PLINK v1.07 ^31^ using the European ancestry-specific dataset available from phase 3 of 1000 genomes as the LD reference^32^. Gene locations were based on GRCh37 (accessed and downloaded via: https://www.cog-genomics.org/static/bin/plink/glist-hg19).

The physical distance threshold for clumping was set to 3000kb, the significance threshold for index SNPs was set to p < 1E-04 and the LD threshold for clumping was r^2^=0.1. Loci were then defined for each index SNP with a p < 5E-08 as the region containing all other variants within the region with r^2^ ≥0.6, in line with previous work^20^.

### Functional Annotation and Gene Set Enrichment Analysis

Functional annotation and gene mapping were performed using FUMA (Functional Mapping and Annotation of Genome-Wide Association Studies; https://fuma.ctglab.nl)^33^. Variants within each of the 19 disorder-differentiating loci were submitted to the SNP2GENE pipeline using default parameters, with eQTL mapping restricted to brain-relevant reference panels: GTEx v8 brain regions (13 regions)^34^ and PsychENCODE^35^. eQTL significance was assessed at FDR < 0.05 within each tissue dataset. Positional mapping assigned genes physically overlapping locus boundaries, whilst eQTL mapping assigned genes for which a locus variant was a significant expression quantitative trait locus in at least one brain tissue. Genes identified by either or both strategies were carried forward as the mapped gene set for downstream analysis.

Gene set enrichment analysis (GSA) was conducted using the GENE2FUNC module within FUMA, applying MAGMA v1.10^36^ for competitive GSA against Gene Ontology (GO) Cellular Component, Biological Process, and Molecular Function collections^37^. Gene sets with fewer than 10 genes were excluded prior to correction. Multiple testing correction was applied separately within each gene set collection using both Benjamini-Hochberg FDR and Bonferroni correction. Tissue expression enrichment was assessed using MAGMA gene expression analysis against GTEx v8 average log_2_(TPM) profiles across 54 tissues, with Bonferroni correction applied across tissues. Due to the limitations of conducting gene set analysis on the small number of loci directly, the full mapped gene set of 102 genes was used as input for the GENE2FUNC analysis.

### Sensitivity Analysis of CC-GWAS Loci using GenomicSEM

Following the CC-GWAS analysis, the shared component of a direct SCZ versus BD GenomicSEM analysis was used to confirm loci that are likely to be truly disorder-specific. The CC-GWAS effect size reflects the difference in SNP association effect sizes between disorders. Consequently, CC-GWAS associated alleles may show concordant risk in both disorders but with a larger effect size in one (convergent alleles). CC-GWAS associations could also reflect alleles conferring risk to only one disorder, or even opposite directions of risk in each of the disorders (divergent alleles). Given the aims of the present study, our primary interest is in divergent alleles. To isolate these, all index SNPs for a locus identified by the CC-GWAS were assessed for shared liability in a published GenomicSEM study of SCZ and BD, and any locus where the index SNP attained a shared association p-value of 1E-05 (a threshold selected to represent nominal association) or less, was then excluded from our final, disorder-differentiating list of CC GWAS loci. The source GWAS for the GenomicSEM analysis were as described above, with the exception that the SCZ summary statistics were restricted to European ancestry only (53,386 cases and 77,258 controls). This is due to the assumption of the GenomicSEM method that summary statistics from all GWAS have been generated on the same ancestry^38^, a requirement that does not apply to the CC-GWAS method.

### SNP-based Heritability and Genetic Correlation Analysis

Disorder-differentiating heritability (observed scale) estimation and genetic correlation analyses were completed using the ‘ldsR’ R package, an R-based implementation of LD score regression^27^. In all instances, the LD reference was the European ancestry-specific data from phase 3 of 1000 Genomes ^32^. SNPs with an INFO score < 0.9 were excluded, and the datasets were trimmed to contain only those alleles present in the third phase of the International HapMap Project ^39^. Phenotypes investigated for genetic correlation were other psychiatric conditions and measures related to cognitive functioning (Figure 4, Supp Table 8). The latter were included because impairments in cognition are noted to be more common in SCZ than BD^40,41^. We also investigated genetic correlation with shared genetic liability across eight psychiatric disorders identified by the PGC ^42^. As a comparator, heritability was also computed using the ‘SumHer’ extension of LDAK^43^. For heritability estimation applied to the CC-GWAS summary statistics, the sample size entered into LDSC and SumHer was set to the combined number of cases across the two input GWAS (67,390 + 41,917 =109,307) rather than the full case plus control totals. This is because CC-GWAS effect sizes reflect differences in allele frequency between case populations, and controls contribute no directional signal to the CC-GWAS statistic. Providing the total case plus control N would artificially inflate the denominator of the LDSC regression, leading to systematic underestimation of disorder-differentiating heritability. Using the case-only N produced a heritability estimate closely concordant with that from an independent GenomicSEM decomposition^44^, providing methodological validation of this approach.

### CC-GWAS PRS Analysis in CardiffCOGs

Using the CC-GWAS results we derived PRS in CardiffCOGS (Cardiff Cognition in Schizophrenia), a cross-sectional SCZ cohort. Consensus diagnoses were based on the Schedules for Clinical Assessment in Neuropsychiatry (SCAN), as well as case note review, according to DSM-IV. The cohort was restricted to individuals with a schizophrenia diagnosis. Additional clinical variables were collected as described previously ^45^. CardiffCOGS received approval from the South-East Wales Research Ethics Committee (07/WSE03/110), and all participants provided written informed consent. Just over a third of participants had treatment resistant schizophrenia (TRS) as defined by a lifetime ever prescription of clozapine (Total SCZ n = 817, TRS = 38.6%), a definition that has been used extensively in studies of treatment resistance^46^. To generate PRS, the CC-GWAS analysis was repeated with a version of the SCZ summary statistics that had CardiffCOGS removed but otherwise processed as above. No individuals from CardiffCOGS were included in any PGC study of BD^24^. PRS were generated using PRSice-2 v2.3.3 ^47^ using non-ambiguous SNPs outside of the MHC region and with an INFO score of > 0.9 and MAF > 0.1. These totalled 3,880,365 SNPs present in CardiffCOGS. A primary p-value threshold of 0.05 was used. As arguments for the PRSice-2 LD clumping procedure, we used clump-kb=250kb, clump-p=1, and clump-r2=0.1, in line with previous PGC work^20^. As a comparator, SCZ and BD PRS were also generated using the same procedure described here, using as training sets the summary statistics inputted to CC-GWAS.

A series of regressions were conducted to assess the relationship between CC-GWAS and eleven SCZ-related phenotypic variables. These included age at onset of psychosis, symptom domains (positive symptoms, negative symptoms of diminished expressivity, negative symptoms of motivation and pleasure, disorganised symptoms, and current cognitive ability), as outlined by Legge and colleagues ^48^, substance misuse, treatment resistance, course of illness, premorbid social / work adjustment, and educational attainment. More detail about these variables and how they were collected can be seen in the **<u>Supplementary Note.</u>** In each case, there were at least 720 individuals with complete phenotypic information in which to test the association. The base model regressed each phenotype on PRS with seven covariates: Principal Components 1 – 5, generated in PLINK v1.07 ^31^, sex, and age at interview. In addition, where appropriate, additional covariates known to be associated with the phenotypic variable under study were added to the model; for substance misuse and educational attainment, year of birth was added, and for course of illness, duration of illness was added. Linear, logistic, or ordinal regression models were used depending on the rating system for each variable.

## Results

### CC-GWAS

In the CC-GWAS analysis (**Figure 1**), 3096 SNPs surpassed GWS, falling within 27 independent loci (**Supp Table 2**). Genomic Inflation factors^49,50^ (λ = 1.201; λ = 1.004), indicated minimal uncontrolled population stratification. Six index SNPs attained p< 1 x 10^-5^ in the GenomicSEM shared component and were excluded, yielding 21 disorder-differentiating loci. In all 6 cases the excluded index SNP showed the same direction of effect in both disorders but a stronger association in SCZ. To account for index SNP fluctuation between analyses, the remaining 21 loci were screened for any SNPs reaching GWS in the BD summary statistics within their boundaries; two further loci met this criterion (both within the MHC region) and were removed. The final set of GWS CC-GWAS results comprised 19 disorder-differentiating loci (**Figure 2**, **Table 1**).

**Figure 1:**
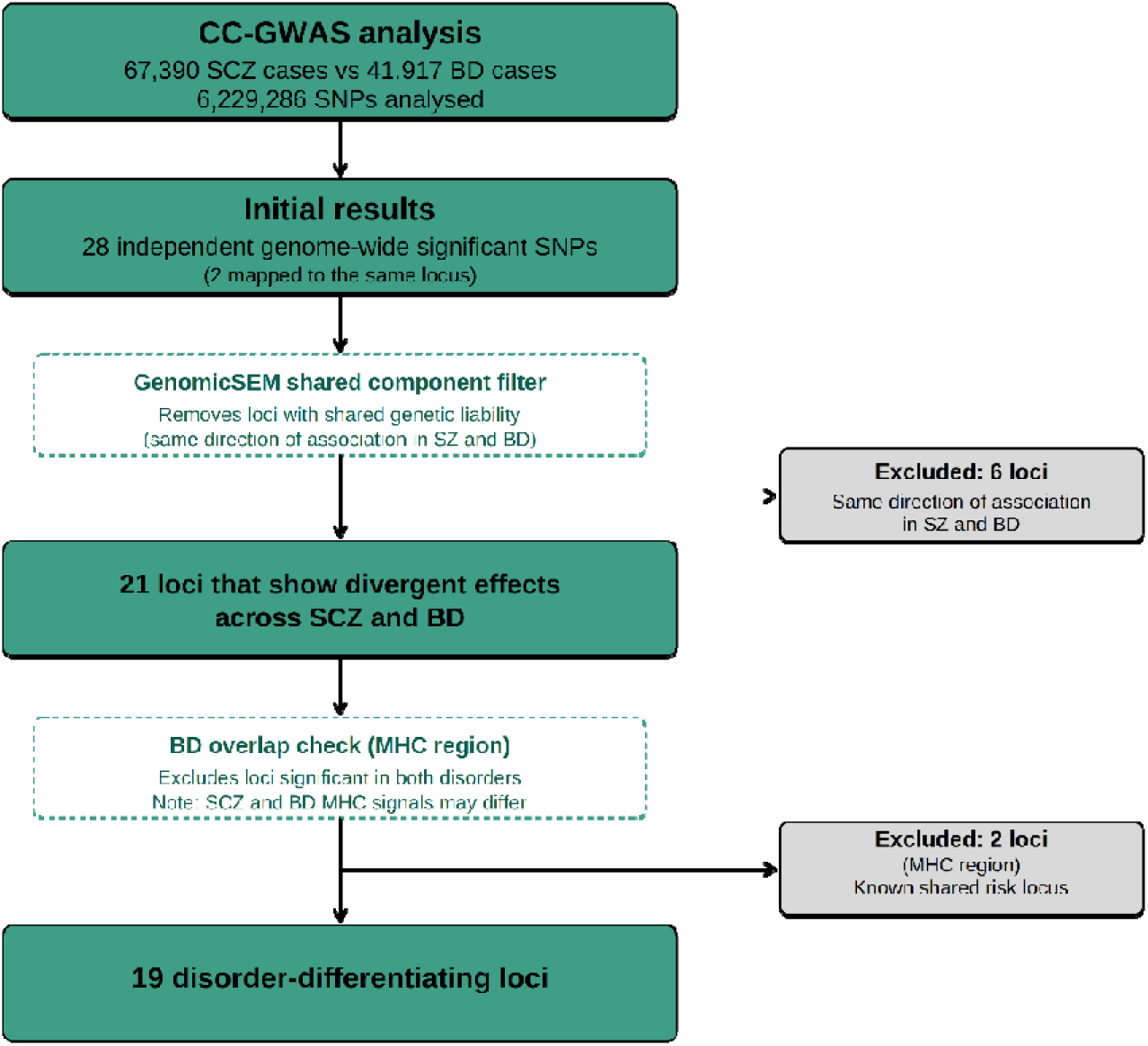
Flow diagram illustrating the progressive refinement from initial CC-GWAS results (27 loci) to the final set of 19 disorder-differentiating loci. Following genome-wide association analysis comparing 67,390 schizophrenia cases with 41,917 bipolar disorder cases, we applied two filtering steps. First, loci showing evidence of shared genetic liability (p<1×10⁻⁵ in the GenomicSEM shared component) were excluded as these represent convergent rather than divergent effects (6 loci removed). Second, two MHC region loci containing multiple g nome-wide significant SNPs in the bipolar disorder GWAS were excluded as these represent known shared risk loci (2 loci removed). The final 19 loci comprised 16 (84%) showing divergent effects (opposite-direction associations between disorders) and 3 showing convergent effects (same direction, larger effect in schizophrenia).

**Figure 2:**
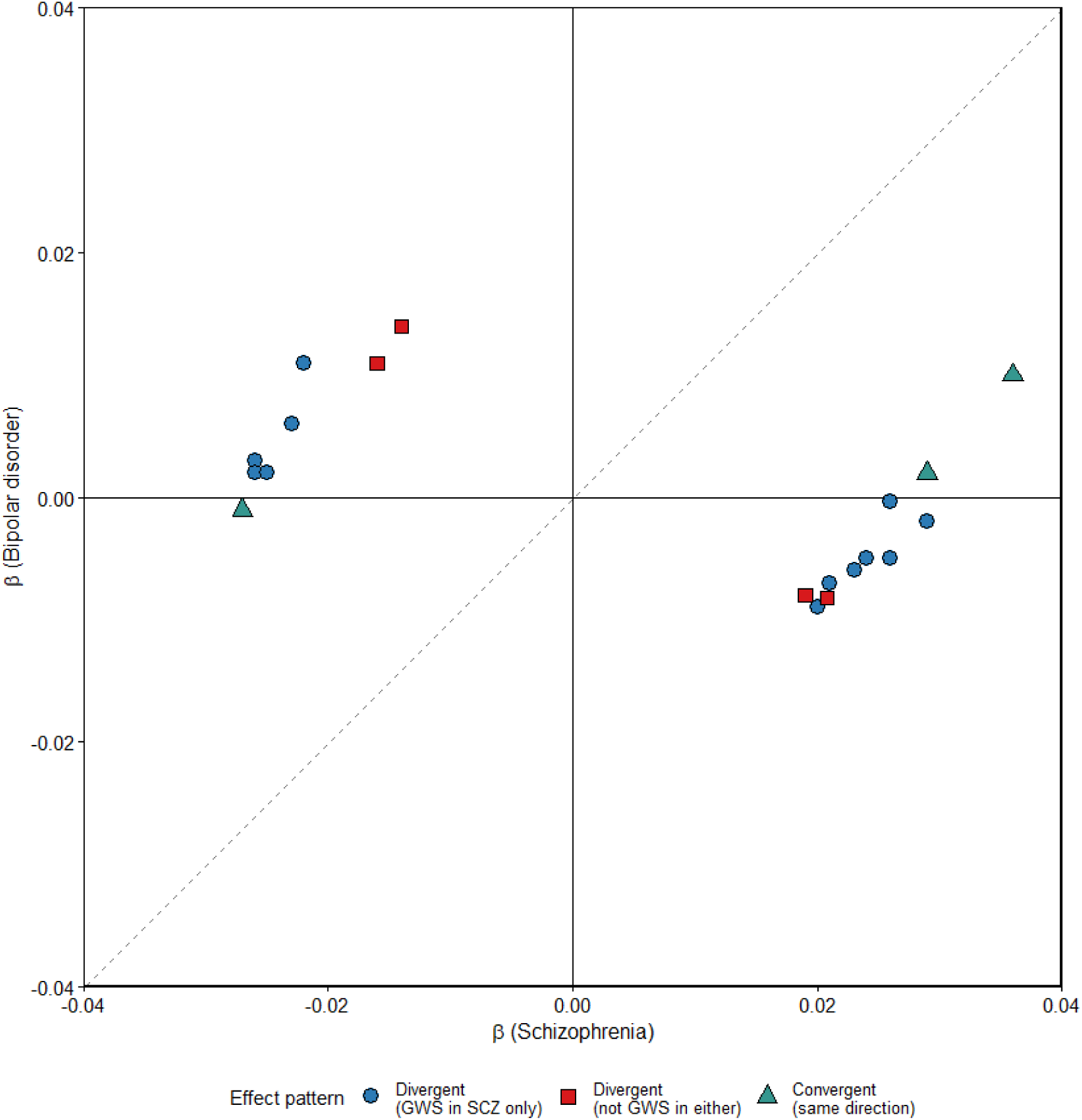
Scatter plot showing effect sizes (β) for the 19 disorder-differentiating loci in schizophrenia (x-axis) versus bipolar disorder (y-axis). Blue circles represent divergent loci with genome-wide significant effects in schizophrenia only (n=12). Red circles represent divergent loci not reaching genome-wide significance in either disorder individually but also showing opposite-direction effects (n=4). Teal circles represent convergent loci showing effects in the same direction but with larger effect sizes in schizophrenia (n=3). The dotted diagonal line indicates β(SCZ) = β(BD); loci in opposite quadrants (top left and bottom right) represent the strongest divergent signals

**Table 1:** The final list of 19 disorder-differentiating loci. SCZ Beta / P refers to the results of these SNPs in the SCZ case:control GWAS, BD to the results in the BD case:control GWAS, and CC-GWAS to the results in the current analysis.

| SNP | CHR | BP | SCZ Beta | SCZ P | BD Beta | BD P | CC-GWAS Beta | CC-GWAS P |
| --- | --- | --- | --- | --- | --- | --- | --- | --- |
| rs9425755 | 1 | 173812639 | 0.020 | 4.36E-08 | -0.009 | 4.6E-03 | 0.015 | 6.01E-10 |
| rs12562967 | 1 | 6797300 | -0.014 | 1.74E-04 | 0.014 | 1.64E-05 | -0.014 | 6.25E-09 |
| rs1822616 | 2 | 55283878 | -0.016 | 2.40E-05 | 0.011 | 4.95E-04 | -0.014 | 2.11E-08 |
| rs1278493 | 3 | 135814009 | -0.026 | 1.35E-12 | 0.003 | 3.37E-01 | -0.016 | 3.99E-10 |
| rs17273111 | 3 | 17855181 | -0.023 | 9.29E-10 | 0.006 | 5.85E-02 | -0.015 | 1.26E-09 |
| rs35746395 | 3 | 181245320 | 0.026 | 1.02E-12 | -0.0003 | 9.3E-01 | 0.014 | 1.34E-08 |
| rs37658 | 7 | 110999503 | 0.026 | 6.06E-12 | -0.005 | 1.71E-01 | 0.016 | 3.60E-10 |
| rs2097942 | 7 | 104725105 | -0.026 | 2.69E-12 | 0.002 | 6.12E-01 | -0.015 | 4.22E-09 |
| rs7838316 | 8 | 27425314 | 0.029 | 3.72E-12 | -0.002 | 5.1E-01 | 0.016 | 1.06E-09 |
| rs10503253 | 8 | 4180844 | -0.025 | 4.37E-11 | 0.002 | 5.35E-01 | -0.014 | 1.75E-08 |
| rs17731 | 10 | 3821561 | -0.027 | 3.76E-13 | -0.001 | 7.57E-01 | -0.014 | 2.79E-08 |
| rs79780963 | 10 | 104952499 | 0.036 | 2.53E-22 | 0.010 | 9.09E-04 | 0.014 | 3.54E-08 |
| rs118031494 | 11 | 133821144 | 0.021 | 1.11E-08 | -0.007 | 2.61E-02 | 0.015 | 2.20E-09 |
| rs3764002 | 12 | 108618630 | -0.022 | 1.65E-09 | 0.011 | 3.30E-04 | -0.018 | 1.27E-12 |
| rs61937595 | 12 | 57682956 | 0.029 | 1.32E-14 | 0.002 | 6.32E-01 | 0.015 | 5.62E-09 |
| rs7048 | 15 | 43620073 | 0.02 | 1.78E-06 | -0.009 | 3.64E-03 | 0.015 | 1.10E-08 |
| rs62039173 | 16 | 4463846 | 0.019 | 1.36E-07 | -0.008 | 1.47E-02 | 0.014 | 9.73E-09 |
| rs62062288 | 17 | 44096553 | 0.023 | 7.53E-09 | -0.006 | 5.97E-02 | 0.015 | 4.01E-09 |
| rs55938136 | 17 | 43798360 | 0.024 | 1.23E-08 | -0.005 | 1.19E-01 | 0.015 | 1.67E-08 |

### Characterisation of Disorder-Differentiating Loci

Of the 19 disorder-differentiating loci, 16 (84%) showed divergent genetic effects (Table 1, Figure 2). In 12 of these, the index SNP was GWS in the SCZ GWAS alone; the remaining 4 divergent loci did not reach GWS in either individual GWAS, demonstrating the enhanced power of CC-GWAS to detect alleles with opposite directions of effects across disorders. Three loci showed convergent effects but substantially attenuated in BD compared to SCZ: Effect alleles were risk-inducing in one (rs17731: SCZ β=-0.027, BD β=-0.001), and protective in the other two (rs79780963: SCZ β=0.036, BD β=0.010; rs61937595: SCZ β=0.029, BD β=0.002). We note none of the 19 loci contained SNPs reading GWS in the BD GWAS, whilst 15 loci contained index SNPs that were GWS in the SCZ GWAS. This is consistent with the greater statistical power of the SCZ input GWAS and the predominantly SCZ-differentiating nature of these signals.

No CC-GWAS locus overlapped any BD locus as identified in the input GWAS, but comparing the CC-GWAS loci with known PGC3-SCZ-associated regions showed these largely colocalise. In terms of physical overlap (using locus boundaries derived from LD clumping), 16/19 (84%) CC-GWAS loci fell entirely within a corresponding SCZ locus, a further 2 had ≥95% overlap, and only 1 showed no overlap with any SCZ locus (rs1822616, divergent, not GWS in either disorder). Assessing the FINEMAP posterior probability (PP) for SCZ captured within each CC-GWAS locus showed full overlap between 12/19 (63%) loci and their corresponding FINEMAP credible sets. In one case the CC-GWAS signal contained a highly credible (“prioritised”) gene, with a single-SNP credible set (ATP2A2). Four other CC-GWAS loci overlapped the fine-mapping credible set of a gene prioritised by the PGC3 SCZ GWAS. For IMMP2L, CSMD1, and KLF6, the entire credible set fell within the CC-GWAS locus boundaries (32/32 SNPs, PP = 0.951; 9/9 SNPs, PP = 0.956; and 3/3 SNPs, PP = 1.000, respectively). For WSCD2, only part of the credible set was captured (3 of 15 SNPs, PP = 0.984 of a total 1.983). WSCD2 was previously flagged in the original CC-GWAS publication as CC-GWAS specific but has since attained GWS in SCZ. This overlap is informative; where correspondence between CC-GWAS and PGC3-SCZ can be established and a prioritised gene has been assigned via fine-mapping, it would be parsimonious to assume CC-GWAS is likely tagging the same underlying association.

#### GTEx Tissue Expression Enrichment

FUMA positional and eQTL mapping across the CC-GWAS summary statistics identified 102 protein-coding genes: 61 by eQTL evidence alone, 19 by positional overlap alone, and 22 by both strategies. eQTL evidence was derived from 13 GTEx v8 brain regions and PsychENCODE. The cerebellum and cerebellar hemisphere contributed the highest number of eQTL entries among GTEx tissues. The predominance of eQTL-only mappings reflects the largely regulatory architecture of psychiatric GWAS signals. All 102 mapped genes were carried forward for GENE2FUNC analysis.

MAGMA gene expression enrichment analysis identified significant enrichment across all 13 GTEx v8 brain regions, all of which survived Bonferroni correction. No non-brain tissue reached statistical significance (Figure 3). The strongest signals were in frontal cortex BA9 (corresponding to a significant portion of the dorsolateral prefrontal cortex, an area strongly implicated in SCZ), brain cortex, and anterior cingulate cortex BA24.

**Figure 3:**
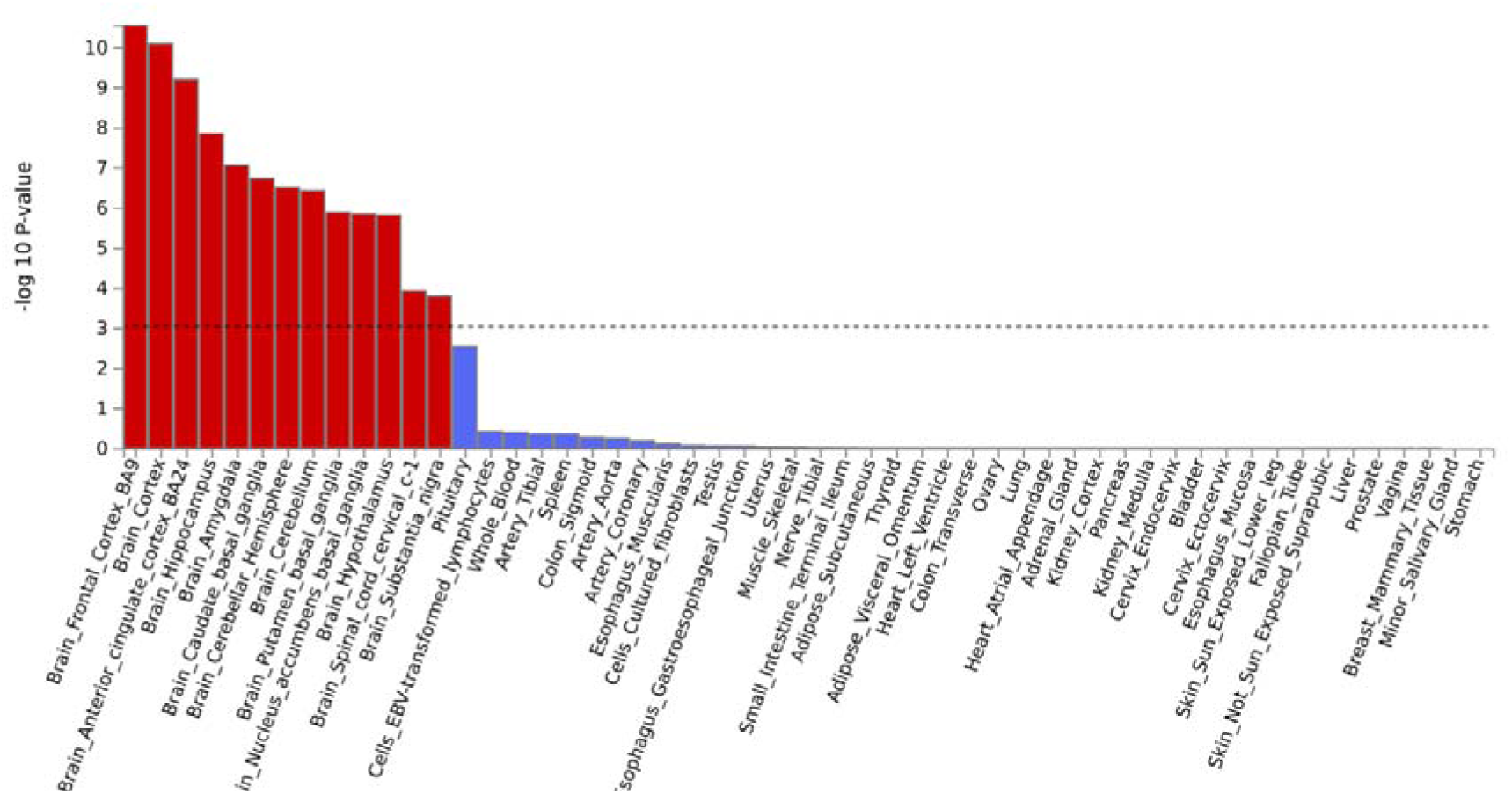
MAGMA gene expression enrichment across GTEx v8 tissues. Bars show −log_10_(p) for gene expression enrichment of the 102 FUMA-mapped genes in each tissue. Red bars = brain tissues; blue bars = non-brain tissues. Dashed line indicates the Bonferroni significance threshold (−log_10_(p) ≈ 3). All 13 brain regions survive Bonferroni correction; no non-brain tissue does. The strongest enrichment is in frontal cortex BA9, general brain cortex, and anterior cingulate cortex BA24.

#### Gene Set Enrichment

MAGMA gene set enrichment analysis (via the GENE2FUNC module, as described in Methods) was conducted across nine gene set annotations. No significant results were observed in “GO:Biological Process” or “GO:Molecular Function”. Within “GO:Cellular Component” (653 sets tested, after excluding sets with fewer than 10 genes), two terms survived Bonferroni correction: NEURON_PROJECTION (N = 1,256 genes, p = 2.51 × 10^-5^, Bonferroni = 0.016) and SYNAPSE (N = 1,363, p = 4.68 × 10^-5^, Bonferroni = 0.031). Full gene set enrichment results are presented in Table 3 and Supplementary Table 9.

#### Heritability and Genetic Correlations

The disorder-differentiating SNP-based heritability was 10.27% (SE=0.01) when calculated via ldsc, and 11.7% (SE=0.01) when estimated via SumHer, both on the observed scale (used because population prevalences are not well-defined for CC-GWAS, as argued in analogous work conducted using GenomicSEM^44^). This is consistent with LDSC guidance that liability scale conversion requires a defined sample and population prevalence for each trait, which CC-GWAS, comparing two case groups directly rather than cases against controls, does not have. The genetic correlation (Rg) between CC-GWAS and SCZ was 0.79 (SE= 0.01), while it was 0.098 (SE= 0.02) with BD, consistent with the predominantly SCZ-differentiating character of the GWS loci.

CC-GWAS was significantly correlated with all traits except for BD2 (Figure 4; Supp Table 7-8). Most strikingly, while both SCZ and BD show positive genetic correlations with educational attainment, CC-GWAS showed a negative correlation, indicating that alleles differentiating SCZ from BD are associated with lower educational attainment. For social deprivation and several cognitive phenotypes (general cognitive performance, numeric reasoning and verbal reasoning), CC-GWAS genetic correlations closely tracked SCZ and were significantly stronger than those for BD. By contrast, correlations with ADHD, BD1, BD2 and AD were much weaker than those of either source GWAS, consistent with these associations being driven by alleles with similar effect sizes on SCZ and BD.

**Figure 4:**
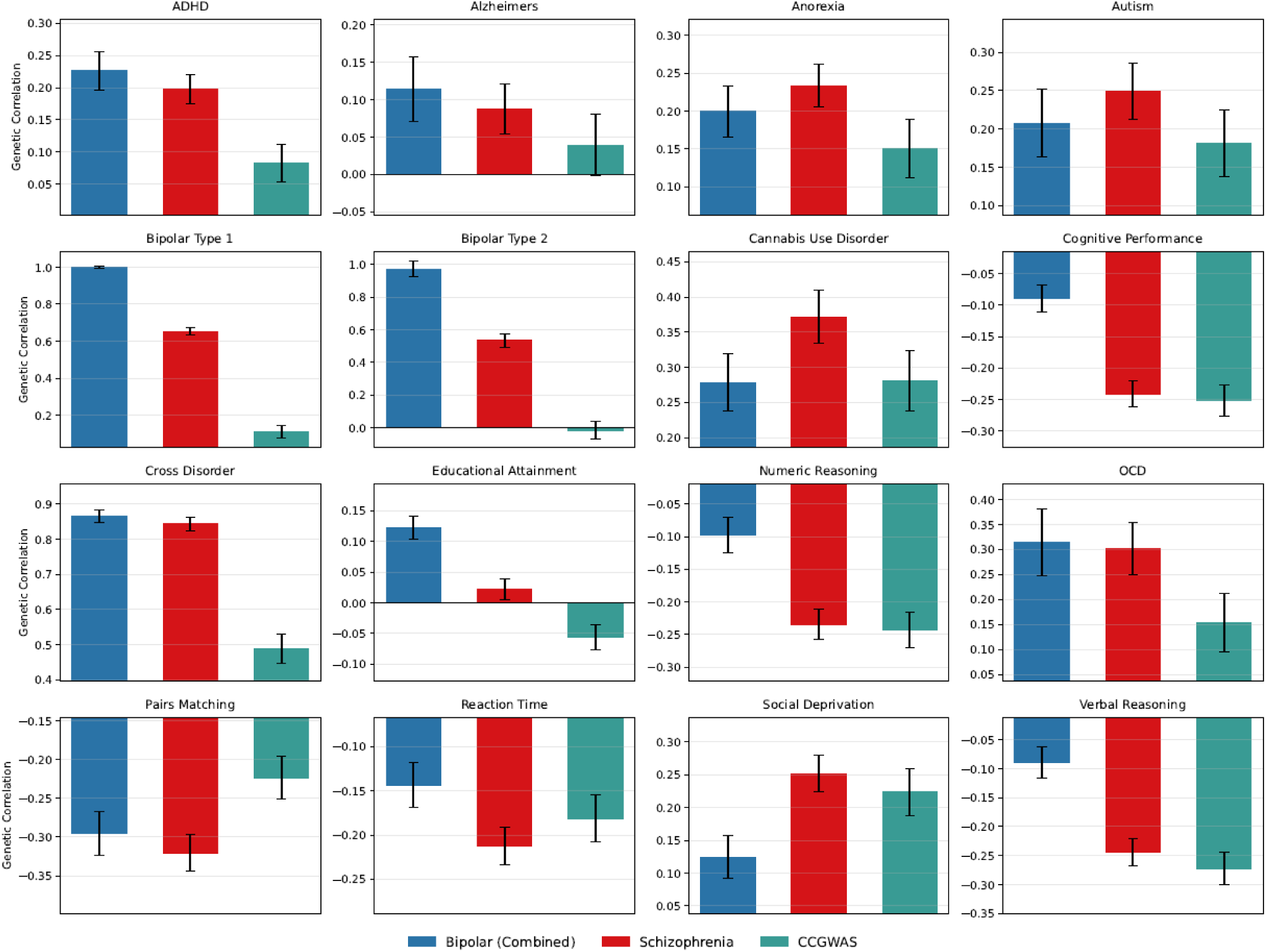
Genetic Correlation Analysis Results for CC-GWAS (Teal), Schizophrenia (Red) and the full Bipolar disorder (Blue) summary statistics with a range of traits, focussing on other major psychiatric disorders, and measures of cognitive performance. Cross-disorder refers to the 2019 paper produced by the cross-disorder working group of the PGC, investigating the pleiotropy shared between the eight major psychiatric disorders^42^

### Polygenic Risk Score Analyses in CardiffCOGS

The CC-GWAS PRS was associated with three clinical phenotypes in CardiffCOGS: younger age at onset of psychosis (β = −0.829, p = 0.009), more severe negative symptoms of diminished expressivity (β = 0.116, p = 0.002), and more severe disorganised symptoms (β = 0.151, p = 0.0001). In each case, the CC-GWAS PRS effect resembled that of the SCZ PRS while the BD PRS showed no significant association, consistent with the alleles influencing these phenotypes being those that differ most between SCZ and BD (**Table 2**; full results in **Supplementary Tables 4-6**).

**Table 2:** Summary table of significant results from the CC-GWAS PRS analysis in CardiffCOGs. ‘Negative symptoms’ refers to negative symptoms of diminished expressivity, ‘disorganised symptoms’ refers to positive thought disorder and inappropriate affect. SCZ = Schizophrenia, BD = Bipolar Disorder. EST = effect size

| PHENOTYPE | CC-GWAS |  | SCZ |  | BD |  |
| --- | --- | --- | --- | --- | --- | --- |
|  | PT_0.05 |  | PT_0.05 |  | PT_0.05 |  |
|  | EST | P | EST | P | EST | P |
| AGE AT ONSET | -0.829 | 0.009 | -1.054 | 0.001 | -0.132 | 0.413 |
| NEGATIVE SYMPTOMS | 0.116 | 0.002 | 0.125 | 0.002 | 0.028 | 0.861 |
| DISORGANISED SYMPTOMS | 0.151 | 0.0001 | 0.2 | 0.0001 | 0.096 | 0.008 |

**Table 3:** MAGMA gene set enrichment results for GO Cellular Component. Results shown for all gene sets surviving FDR < 0.05 within-collection correction. Bold rows indicate sets additionally surviving Bonferroni correction. N genes = number of genes in the gene set; Beta = MAGMA regression coefficient; Beta (std) = standardised beta; FDR = Benjamini-Hochberg false discovery rate; Bonf = Bonferroni-corrected p-value, both applied within each gene set collection separately. Gene sets with fewer than 10 genes were excluded prior to correction. VGCC=Voltage Gated Calcium Channels

| Gene set | N<br>genes | Beta | Beta<br>(std) | P-value | FDR | Bonf |
| --- | --- | --- | --- | --- | --- | --- |
| **NEURON_PROJECTION** | 1256 | 0.1161 | 0.0289 | 2.51E-05 | 1.53E-02 | 1.64E-02 |
| **SYNAPSE** | 1363 | 0.1074 | 0.0277 | 4.68E-05 | 1.52E-02 | 3.06E-02 |
| **CHROMOSOME** | 1726 | 0.0894 | 0.0257 | 1.33E-04 | 2.48E-02 | 8.66E-02 |
| **DENDRITIC_TREE** | 572 | 0.1475 | 0.0252 | 1.68E-04 | 2.48E-02 | 1.10E-01 |
| **L_TYPE_VGCC_COMPLEX** | 12 | 1.0476 | 0.0263 | 1.9E-04 | 2.48E-02 | 1.24E-01 |
| **CHROMATIN** | 1190 | 0.1018 | 0.0247 | 2.83E-02 | 2.83E-02 | 1.7E-01 |
| **NUCLEAR_BODY** | 754 | 0.1191 | 0.0233 | 3.57E-02 | 3.57E-02 | 2.5E-01 |
| **AXON** | 600 | 0.1348 | 0.0236 | 3.62E-02 | 3.62E-02 | 2.9E-01 |

## Discussion

Using CC-GWAS to analyse 67,390 SCZ cases and 41,917 BD cases, we identified 19 genome-wide significant loci that differentiate the two disorders. Sixteen of these loci showed divergent genetic effects, whereby risk alleles have opposite directions of association between SCZ and BD; only three were concordant. This pattern indicates that, despite their strong genetic correlation, the loci differentiating SCZ and BD predominantly reflect qualitative differences in genetic architecture (opposite-direction effects) rather than purely quantitative differences in effect magnitude (same-direction effects). The loci with the strongest divergent effects converged on genes involved in neurodevelopment, particularly neuronal structure, and axon extension, and synaptic function, consistent with longstanding hypotheses that SCZ has a more pronounced neurodevelopmental component than BD^52^.

The three convergent loci, in which the same allele conferred risk for both disorders but with substantially attenuated effects in BD, are also informative. Rather than identifying biology that is unique to one disorder, these loci likely index shared molecular pathways for which SCZ-specific mechanisms amplify or modulate liability. That these loci survived the GenomicSEM shared component filter suggests the BD effect sizes are genuinely small relative to those in SCZ, rather than reflecting fully shared signal. Their retention within the disorder-differentiating set therefore highlights regions where quantitative rather than qualitative genetic differences between disorders may operate, a distinction that is relevant for understanding the dimensional nature of psychosis liability. This may indicate their relevance for phenotypes that operate across the two disorders.

Of particular note are the four loci that reached genome-wide significance in the CC-GWAS despite not attaining significance in either individual disorder case-control GWAS. These signals (index SNPs: rs12562967, rs1822616, rs7048, rs62039173) appear in our results due to their opposite directions of effect across the two disorders^53^. None of these loci reached genome-wide significance in the individual SCZ or BD GWAS and therefore were not included in the PGC3 fine-mapping analysis. They offer the clearest demonstration of the added value of CC-GWAS as a discovery tool and warrant particular attention in future functional follow-up studies.

Functional annotation of the summary statistics via FUMA identified 102 protein-coding genes through positional and eQTL mapping against brain-relevant reference panels, providing a broader biological context than locus-level assignment alone. Gene expression enrichment analysis confirmed that this mapped gene set is preferentially expressed in brain tissue, with significant enrichment across all 13 GTEx v8 brain regions and no significant enrichment in any peripheral tissue. This specificity is notable given the case-case design, which conditions out the substantial shared genetic liability between SCZ and BD. The disorder-differentiating signal retains a distinctly brain-specific expression profile, consistent with mechanisms operating within brain tissue rather than being distributed across peripheral systems.

Gene set enrichment analysis implicated neuronal compartment and synaptic annotations as the most robust biological themes distinguishing SCZ from BD. Two GO Cellular Component terms survived Bonferroni correction: GOCC_NEURON_PROJECTION and GOCC_SYNAPSE. These findings are consistent with established differences in synaptic connectivity, dendritic architecture, and neurotransmitter receptor expression between SCZ and BD, and align with the broader neurodevelopmental hypothesis of SCZ^52,54^. A nominally significant signal was also observed for the L-type voltage-gated calcium channel gene set (FDR < 0.05), of potential interest given the well-replicated involvement of L-type voltage-gated calcium channels in cross-disorder psychiatric GWAS signals^55^.

We note the results from one locus should be considered with caution. The chromosome 17q21.31 region shows pleiotropic associations across a wide range of traits including Parkinson’s disease^56^, Alzheimer’s disease^57^, and alcohol use^58^. It contributes two independent signals and 2,078 SNPs to our CC-GWAS results and spans the well-known MAPT H1/H2 inversion haplotype^59^.

Overall, our CC-GWAS summary statistics had detectable heritability (10.2%–11.7%). Genetic correlations broadly mirrored those of SCZ, with positive associations across several psychiatric phenotypes and negative associations with cognitive performance.

The sole exception to this convergence was educational attainment, where SCZ shows a small positive genetic correlation, but CC-GWAS shows a negative one. This was consistent with a recent GenomicSEM decomposition in which the SCZ-differentiating signal associated with lower educational attainment and poorer cognition^44^. Associations between SCZ and educational attainment have been inconsistent in the literature, and both positive^60^ and negative^61^ correlations have been reported. This inconsistency may be at least partly attributable to differences in phenotype definitions across cohorts^62^. However, it could still be informative: the positive genetic correlation between SCZ liability and educational attainment is likely driven by the large proportion of SCZ genetic architecture shared with BD, given that BD shows a consistently positive genetic correlation with educational attainment. The subset of variants conferring SCZ-specific risk appears instead to be enriched for pathways that impair cognitive development, producing the negative association observed in the CC-GWAS. This interpretation is consistent with Richards et al. (2025)^44^ and suggests that prior reports in the literature may have been affected, at least in part, by how the shared SCZ-BD liability varied across study samples.

PRS derived from the CC-GWAS loci showed several associations in the CardiffCOGS schizophrenia cohort. Higher CC-GWAS PRS was associated with earlier illness onset, as was SCZ PRS but not BD PRS. Younger age at onset in schizophrenia associates with greater symptom severity and poorer outcomes^63^, and seeing this pattern in SCZ PRS is consistent with the idea that greater inherited liability likely accelerates the emergence of illness. The relationship also holding for CC-GWAS (which indexes liability not shared with BD) reinforces the interpretation that SCZ-differentiating variants might influence early neurodevelopmental trajectories.

Both SCZ PRS and CC-GWAS PRS, but not BD PRS, were associated with more severe negative symptoms of diminished expressivity (flattened facial affect and poverty of speech^64,65^), suggesting that genetic variation unique to SCZ contributes disproportionately to symptoms most closely tied to neurodevelopmental impairment. Negative symptoms, particularly when distinguished from depressive symptoms, are considerably more prevalent in SCZ than BD, raising the possibility that part of the SCZ-specific genetic signal reflects liability to this symptom domain specifically. Indeed, the severity of negative symptoms has previously been linked to exposures associated with disrupted neural development, such as prenatal infection and maternal deprivation^66^. A related hypothesis is that liability shared with BD may load more heavily on affective or episodic features of illness, rather than on cognitive and negative symptoms, though we did not observe significant results for these symptom domains in our analyses.

For disorganised symptoms (positive thought disorder and inappropriate affect), all three PRS showed positive association, with effect sizes for SCZ PRS and CC-GWAS PRS both exceeding that of BD PRS. This pattern suggests that the relevant genetic liability is distributed across the psychosis spectrum, with SCZ-specific variants exerting a stronger but not exclusive effect: a difference in degree rather than in kind.

### Replication of Previous Results

The paper outlining the CC-GWAS method^19^ reported an analysis of SCZ and BD based on the largest publicly available case-control GWAS for each disorder at the time of publication ^20,21^. As both GWAS were PGC studies, there is a substantial sample overlap with the GWAS used here. Methodologically, our use of CC-GWAS only differed in the parameters for the calculation of FST_causal_, which were tailored to our input datasets. Thus, we consider our current results as a “narrow replication”^67^ of this earlier work, where the sample overlap allows us to assess how CC-GWAS results change with the increases in sample size and power of the SCZ and BD GWAS.

Of the index SNPs reported in the earlier paper (loci were not defined), 3 of the 12 were significant in this CC-GWAS analysis. One of these (corresponding to the WSCD2 SCZ locus) was reported as a “CC-GWAS specific signal” in the earlier paper, which meant that neither the SNP nor close LD proxies (r^2^>0.8) were detected as GWS in the input case-control GWAS results. However, the index SNP (rs3764002) is now significant in the SCZ GWAS used for our analysis^22^. This is another indication that CC-GWAS allows for the earlier detection of some signals that, through increases in sample size and statistical power, will eventually become GWS in disorder-specific analyses.

In a direct case-case GWAS of 20,129 individuals with BD and 33,426 individuals with SCZ conducted in 2018 by Ruderfer et al., two loci were identified with divergent effects on SCZ and BD; rs56355601 within DARS2, and rs200005157 within ARFGEF2 ^15^. In both, MAF was higher in BD cases than in SCZ cases. In the present study, the DARS2 association replicated, but rs56355601 was more significantly associated with SCZ rather than BD. Regarding ARFGEF2, the index SNP from the earlier study was absent from the CC-GWAS dataset, and the minimum P-value observed was 7.5×10^-6^. This discrepancy may reflect differences in power across the two studies or differences in ancestral composition, as the Ruderfer et al. analysis was restricted to European ancestry cases.

Byrne and colleagues, using mtCOJO to generate SCZ summary statistics conditioned on four other psychiatric disorders, identified 15 of 130 SCZ loci from the input SCZ GWAS^20^ as particularly SCZ-specific^68^. Only one of these (WSCD2) overlaps with a CC-GWAS locus in the present analysis, which is further evidence that this is a SCZ-specific gene of particular interest. The discrepancy most likely reflects different input GWAS as well as the conditioning on multiple disorders (BD, MDD, ADHD and ASD).

Compared with a direct case-case GWAS requiring individual-level genotype data, CC-GWAS can leverage large-scale GWAS summary statistics that are generally more readily accessible to the scientific community. It is also less sensitive to allele frequency differences caused by ancestral or genotyping array variability, and this allowed for retaining East Asian cohorts in our analysis. By deriving case-case associations from matched case-control summary statistics rather than comparing case groups directly, CC-GWAS accounts for population stratification through the respective control groups. The method also includes a formal filtering step for potential false positive associations caused by so-called “stress test” SNPs, a source of artefactual divergence that can arise from sampling variance or imputation differences in the input GWAS.

### Limitations

There are several limitations to consider. First, the identified loci were primarily SCZ-differentiating, likely because the SCZ input GWAS has greater statistical power and more GWS loci than the BD input GWAS. This imbalance limits the discoverability of true bipolar-specific effects and highlights the need for future work leveraging larger, phenotypically homogeneous BD cohorts. Although a larger BD GWAS has been published since our analyses were conducted^25^, we used the 2021 GWAS due to concerns about phenotypic heterogeneity (see **Methods**).

Second, both SCZ and BD are clinically heterogeneous disorders with variable presentations, courses, and outcomes. Our analysis treats each as a single diagnostic category, which may obscure genetic effects specific to particular subtypes or symptom profiles. For instance, individuals with schizoaffective disorder were included in the SCZ GWAS, and BD cases comprised both type 1 and type 2 diagnoses. The disorder-differentiating signals we identify may relate more strongly to specific clinical presentations within each broad diagnostic category or symptom domains that cross disorders but are more frequent in SCZ (e.g. in the case of negative and disorganised symptoms).

Future studies examining genetic differentiation across finer-grained clinical dimensions or subtypes may reveal additional insights into the biological basis of diagnostic boundaries.

Third, although East Asian cohorts were included in the input dataset we used for SCZ, the current analysis is nevertheless predominantly based on data from individuals of European ancestry, limiting generalisability. As CC-GWAS can accommodate ancestral diversity in its input datasets, replication in more globally representative samples is a natural next step.

Finally, the CC-GWAS method is sensitive to sample size imbalances and assumptions about trait independence, and may be less powerful than an equivalent direct case-case analysis when the control populations used in the original GWAS differ systematically. Nonetheless, its ability to uncover novel loci with disorder-specific relevance, combined with its use of publicly available summary statistics rather than individual-level data, remains a valuable contribution to psychiatric genetics.

Together, our findings demonstrate that genuine divergent genetic effects exist between SCZ and BD beyond their substantial shared liability. The predominance of divergent over convergent effects (84% versus 16%), the convergence of divergent loci on neuronal and synaptic processes, and the associations with cognitive impairment and early onset collectively support the hypothesis that SCZ-specific genetic risk is characterised by greater neurodevelopmental disruption. The identified loci and biological pathways provide concrete targets for mechanistic research into what distinguishes these disorders at a molecular level, as well as a foundation for developing disorder-specific biomarkers and therapeutic strategies.

## Data Availability

To comply with the ethical and regulatory framework of the CardiffCOGS cohort, access to individual-level data requires a collaboration agreement with Cardiff University. Requests to access deidentified datasets, data dictionaries, and other summaries from the CardiffCOGS cohort should be directed to the corresponding authors

