## Supplementary Note for "Identifying and Characterising Common Genetic Differences in Schizophrenia and Bipolar Disorder"

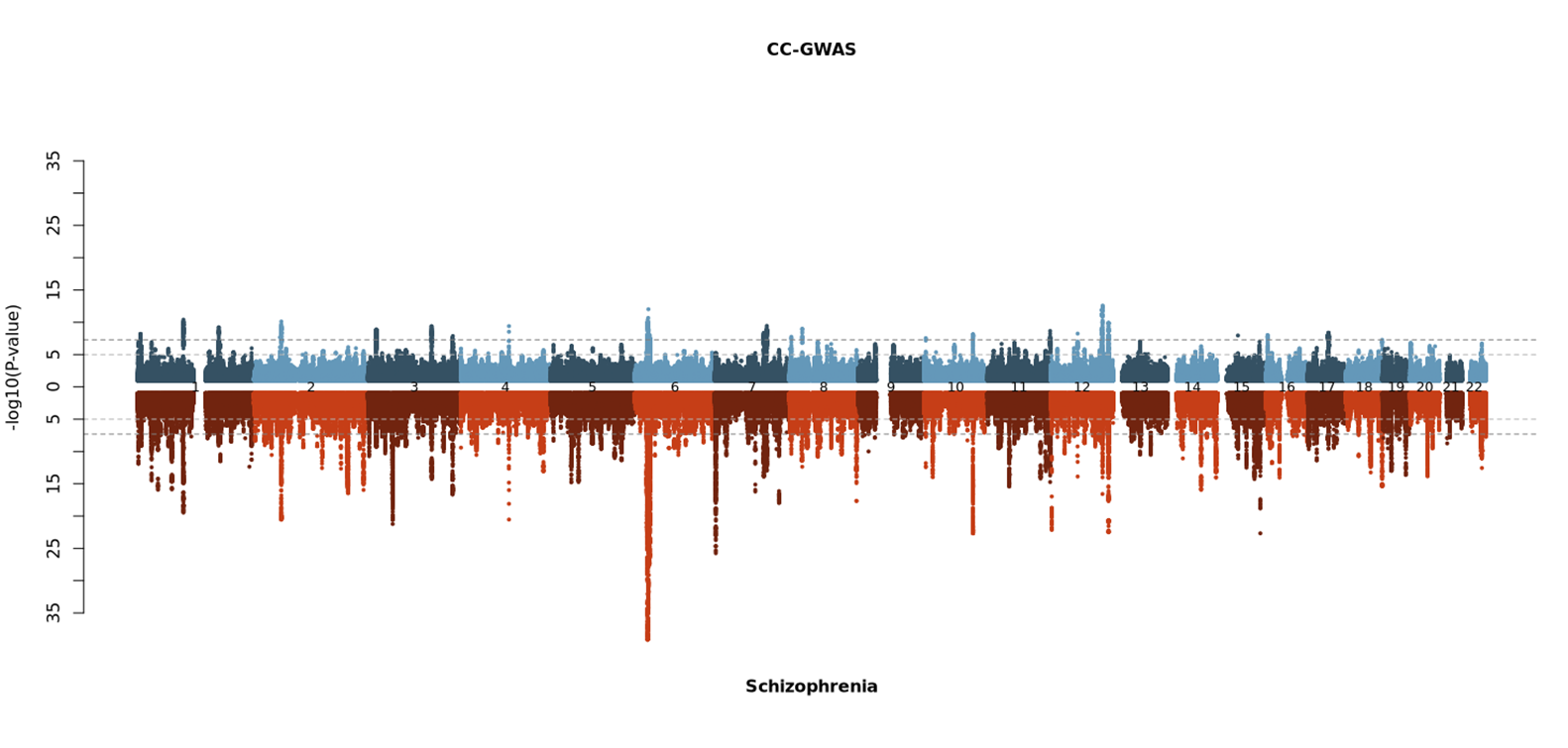

Figure 1: Miami Plot of CC-GWAS summary statistcs versus Schizophrenia PGC3 case:control

### Supplementary Phenotype Description

Below is a series of descriptions from the phenotypic variables investigated in the PRS analysis of this paper. They are taken directly from the interview ratings guide used by interviewers at the time of participant ascertainment

**Measures of Premorbid Function:**

**Poor Work Adjustment:**

Refers to work history before onset of illness. It should be scored if the patient was unable to keep any job for more than 6 months, had a history of frequent changes of job or was only able to sustain a job well below that expected by his educational level or training at time of first psychiatric contact. Also score positively for a persistently very poor standard of housework (housewives) and badly failing to keep up with studies (students). (0,1)

**Poor Premorbid Social Adjustment:**

Patient found difficulty entering or maintaining normal social relationships, showed persistent social isolation, withdrawal or maintained solitary interests prior to onset of psychotic symptoms. (0,1)

If other relevant information is absent, a patient having had no friends at school or only one casual friend is rated ‘1’. Patients who had any more casual friends or any good friends are rated ‘0’.

**Course of Disorder:**

1= Single episode with good recovery

2= Multiple episodes with good recovery between

3= Multiple episodes with partial recovery between

4= Continuous chronic illness

5= Continuous chronic illness with deterioration (nb score this item in hierarchical fashion, eg, if patient's course in past rated '2',but for the time-period now being considered it rates '4', then the correct rating is '4'.)

Choosing between ratings of ‘4’ and ‘5’ - most patients with incapacitating chronic schizophrenia are rated as ‘4’ unless it is clear that they have gradually deteriorated in terms of severity of symptoms from the start of their illness to the present.

Choosing between ratings of ‘3’ and ‘4’ - if the patient appears to have had active positive psychotic symptoms or incapacitating symptoms for the majority of the illness duration, rate as ‘4’. If they have had these symptoms for the minority of the duration of the illness in episodes with residual symptoms between, they can be rated as ‘3’.

Rate ‘2’ if the patient resumes pre-morbid function between episodes.

RATE TO ONE DECIMAL PLACE IF NECESSARY, e.g., 2.5 suggests a course of disorder that is exactly midway between a rating of 2 and a rating of 3. 2.2 suggests a course of disorder that is part-way between a rating of 2 and a rating of 3 but is closer to a rating of 2 than a rating of 3.

**Treatment Resistance**

Determined via use of clozapine (Clozapine_Ever=YES) and / or OPCRIT Item 89=0:

Psychotic symptoms respond to neuroleptics.

Rate globally over total period. Score positively if illness appears to respond to any type of neuroleptics (depot or oral) or if relapse occurs when medication is stopped. (0, 1)

Rate as ‘1’ if a substantial improvement in psychotic symptoms either subjectively or according to notes or informants. Rate as ‘0’ if patient on Clozapine because of treatment resistance. Rate as default 9 if no information in case notes.

**Substance Abuse**

**Cannabinoids ever regular?**

0	No

1	Yes

9	Unknown

Nb. ‘Regular use’ means persistently for one month or repeatedly within one year (i.e. at least once a week for at least 6 months of the year)

**Other non-Prescription Drugs**

Unspecified drugs ever regular?

0	No

1	Yes

9	Unknown

Nb. ‘Regular use’ means persistently for one month or repeatedly within one year (i.e. at least once a week for at least 6 months of the year)

**Educational Attainment**

This variable expresses the highest level of education reached by the participant. This is a numeric variable ranging from 0 to 6 and includes 9. The encoding is as follows:

0 = None

1 = 11+

2 = CSE, Certificate of Secondary Education (awarded from 1965 to 1987). Designed for people not pursuing the GCE O-levels.

3 = O-level/GCSE

4 = A-level

5 = Degree

6 = Post-graduate degree

9 = Unknown
